# Applied Ontologies for Global Health Surveillance and Pandemic Intelligence

**DOI:** 10.1101/2020.10.17.20214460

**Authors:** Christopher J. O. Baker, Mohammad Sadnan Al Manir, Jon Hael Brenas, Kate Zinszer, Arash Shaban-Nejad

**Author notes:** Corresponding Authors: Arash Shaban-Nejad, PhD, MPH, The University of Tennessee Health Science Center (UTHSC) - Oak Ridge National Laboratory (ORNL) Center for Biomedical Informatics, Department of Pediatrics, College of Medicine, 50 N Dunlap Street, R492, Memphis, Tennessee 38103, United States, and Christopher J. O. Baker PhD Department of Computer Science, University of New Brunswick, Canada 100 Tucker Park Rd, Saint John, NB E2K 5E2.

## Abstract

Global health surveillance and pandemic intelligence rely on the systematic collection and integration of data from diverse distributed and heterogeneous sources at various levels of granularity. These sources include data from multiple disciplines represented in different formats, languages, and structures posing significant integration challenges This article provides an overview of challenges in data driven surveillance. Using Malaria surveillance as a use case we highlight the contribution made by emerging semantic data federation technologies that offer enhanced interoperability, interpretability and explainability through the adoption of ontologies. The paper concludes with a focus on the relevance of these technologies for ongoing pandemic preparedness initiatives.

## 1 Introduction

Whereas Health surveillance has always been an essential activity, the recent global health crisis caused by COVID-19 has highlighted our dependence on bespoke surveillance infrastructures that provide support for decisions of considerable gravity. Core to the success of these endeavors is the coordination of multiple data sources, however, many challenges related to data management remain unresolved and limit the insight we can gain from data-driven surveillance. These challenges stem from incumbent infrastructures where collected data is stored in distributed heterogeneous siloed information systems without enabling metadata or standardization, posing interoperability and data integration challenges [1]. The absence of interoperability results in poor coordination between surveillance systems which are known to be rigid [2], leading stakeholders to have limited confidence in any insights derived from the aggregated datasets [3]. These barriers delay the generation of key insights that are needed to support decision making and to plan appropriate and timely interventions. Specifically for decision-makers, there is also an intelligibility problem, which requires a transparent understanding of sources of data and data processing activities to generate trustable insights. Here, surveillance systems need to enable explanations based on provenance annotations. The requirement for explainability becomes increasingly important given the diversity of data sources being harvested in disease surveillance. Indeed, an increasingly broad array of stakeholders now contributes vast sensor data from new devices and text from social media platforms, augmenting the complexity of sources and data structures.

In the case of malaria, it has been reported that few information systems can comprehensively collect, store, analyze data, and provide feedback based on real-time information [4]. Additionally, concurrency control issues can emerge, where data entered from field stations are recorded centrally but are not immediately reflected at the field level, [5-7]. Developers and users trying to gain remote access to data with web services have also identified that they are inflexible for reuse and there is a lack of standardization. In fully deployed systems, the resolution of spatiotemporal data [8] is often limited in scope, e.g. not to the level of individual households, nor is it possible to extrapolate trends across geographic borders. Consequently, the types of surveillance queries that can be run to derive actionable knowledge are relatively rigid and reporting tools cannot support ad hoc queries without significant redevelopment. A recent WHO technical strategy document [9] stated that malaria surveillance mechanisms designed to facilitate interoperable data integration from distributed data silos are lacking.

These types of challenges, interoperability, interpretability, and explainability, have motivated computer scientists to consider how the provision data for a wide range of stakeholders leading to the introduction of a set of guidelines seeking to ensure that published data are findable, accessible, interoperable, and reusable [10_5_], albeit no specific technical solutions are proposed or mandated.

Broadly speaking, interoperability can be understood as the provision of common interfaces between divergent computer systems to ensure seamless access to data and services. It can include enriching data or services with context, unambiguous meaning, and provenance in a standardized syntax or format in support of data exchange and reuse. Practically, it means multiple users can mutually access and reuse data without having to store it locally or reformat the data on import, knowing that the integrity and meaning (semantics) of the data has been maintained since its creation to its application and reuse. Provisioning interoperability is addressed by the mapping of data to standardized vocabularies or terms in ontologies [11]. Ontologies are representations of domain knowledge using concepts, relations, and complex logical rules or axioms. Interoperability can be achieved by mapping data sets to the same ontology terms or vocabularies to ensure they can be regarded as having the same meanings.

Such representations of domain knowledge and metadata also underpin the explainability of decision support in a surveillance platform where the reasoning behind the decision can be made transparent to the end-users. In order to achieve this transparency, the data resources supporting a decision need to be verified, preferably in real-time. One method to verify the resources used to achieve the decision is by resolving all their locations from their URIs. Once resolved, they lead to the locations of the actual content or the associated metadata based on the access policy. In a service-based surveillance system [12], resources typically include the input data, and services, workflows, software, and scripts which produce the output data. Various approaches to verification also referred to as *provenance*, have been investigated and tried in the context of data [13], software [14], and workflow [15, 16]. A standard for vocabularies to represent provenance information in a formalized way is the W3C PROV Ontology (PROV-O)[17] which can be used as a vehicle to formalize explainability.

## 2 Disease Surveillance Ontologies

To be effective, disease surveillance ontologies [18, 19, 20, 21] must cover a range of perspectives including vector biology, etiology, transmission, pathogenesis, diagnosis, prevention, and treatment. Depending on the intended use of the ontology, these perspectives have been represented in different ways leading to ontologies of different maturity, expressiveness, and fit-for-purpose. Whereas a detailed review of these would be beyond the scope of this article, here we briefly review a small sample of ontologies primarily to illustrate the breadth and scope of the domain knowledge that needs to be represented to support surveillance interoperable surveillance infrastructures.

For vector-borne diseases, the Vector Surveillance and Management Ontology (VSMO) [18] focuses on arthropod vectors and vector-borne pathogens specific to domestic animals and humans as well as the corresponding surveillance data management systems, or decision support systems. A core feature of the VSMO is the has_vector relation, linking arthropod species (vectors) to pathogenic microorganisms.

The Infectious Disease Ontology-Malaria (IDOMAL) [19] is an extension of the infectious disease ontology (IDO) [22] that includes broad coverage of malaria including clinical manifestations, therapeutic approaches, epidemiology, vector biology, and insecticide resistance (IR). Vector physiology is modeled from the perspective of processes related to transmission, particularly interactions between the vector and the vertebrate host of Plasmodium, as well as the vector and Plasmodium itself. This extends to behavioral parameters such as host-seeking or blood meal-related processes.

Mosquito Insecticide Resistance Ontology is an application ontology for entities related to insecticide resistance in mosquitoes. MIRO [20] was designed to support IRbase, a dedicated resource for storing data on insecticide resistance in mosquito populations. It focuses on archiving information on geolocations, mosquito populations as the main vectors of diseases (dengue, filariasis, malaria, yellow fever) types of assays performed to assess types and levels of insecticide resistance to support the design of interventions.

Animal Health Surveillance Ontology (AHSO) [21] is an excellent example of this, designed to support decision making based on data that were collected for alternative purposes, including clinical records, laboratory findings, or slaughter inspection data. In this case, herd information is collected along the entire cycle of animal production, even in the absence of disease events. The targeted surveillance analytics makes use of all recorded observations such as a disease occurrence, births, or product yield (dairy or livestock). Ontologies that support surveillance analyses and automation of tasks translating data into actionable information must accommodate the production system, the nature of observation, and the context in which the data was recorded. AHSO represents definitions of syndromes and models observations that relate to health events at specific moments in time but not the actual health events. The ontology has three main levels: sample, observations, and observational context, for instance, a clinical observation, or surveillance sampling activity. A health event is modeled as an abstract concept with undefined boundaries in time, space, or population units. It is assumed that several observations are derived from a health event and recorded in one or more databases [21].

In general, the adoption of ontology models by users other than the ontologists that built them can depend on many factors related to target goals and purpose. The design of an ontology, so that it is fit for purpose, can vary greatly, and ontologies designed for different purposes by different communities generally result in different ontologies, both in terms of scope and structure, which can occur for even the same subject matter.

Deciding whether a domain ontology built for any of these purposes and auxiliary activities can be reused is daunting and requires considerable expertise and time-investment. For these reasons, the reuse and adoption of any given ontology is generally a slow process requiring a full evaluation of what aspects of an ontology can be useful in a new context. Sometimes it is easier to start a new ontology and then subsequently do a mapping to any related ontologies that may exist. Where ontologies or parts of ontologies have been imported to new ontologies this can be identified by reviewing ontology files for imports with different URLs to reveal which parts of an ontology are from other conceptualizations. Some studies have sought to elucidate such artifacts [39] with varying degrees of success.

To further highlight the challenges of reuse we list here some common motivations for ontology development; (i) to share a common understanding of information, (ii) to enable reuse of knowledge through explicit representation of knowledge and formal reasoning, (iii) the derivation of further insights in a domain aka knowledge discovery, (iv) to enable reasoning and quality control (i.e. revealing inconsistencies and insatisfiabilities), and (v) to improve reusability, maintenance, versioning and change management. The precise manifestation of these goals can be quite technical and here we point primarily to the classification of ontologies to identify inconsistencies in a knowledge representation about a domain [23], classification of instance data based on formal axioms or rules in a domain ontology [24, 25], authoring of knowledge graphs using ontologies as a reference model [26], ontology-based data access [27], and the use of ontology terms for authoring of semantic web services [28]. All of these are specific cases that leverage more than the primary formal conceptualization of a domain built for the sake of knowledge sharing. Overall the maturity of the model and its design and purpose are limiting factors for reuse. One good example of an ontology that has been well cited and adopted is the Semantic science Integrated Ontology (SIO) [29] which provides a simple, integrated upper-level ontology (types, relations) for consistent knowledge representation across physical, processual, and informational entities. It is broadly adopted in the life sciences because of its design and relevance to many use cases.

## 3 Disease Surveillance tasks

Surveillance is an activity that involves a series of tasks; monitoring and harvesting of data, analysis of data to review the disease trends followed by the design of targeted interventions, their implementation, and evaluation. To be effective, surveillance is an iterative lifecycle of tasks and activities with the goal of harvesting actionable data. What makes it challenging is that surveillance practitioners need to be able to obtain custom views of target data and decision parameters in a timely and non-arbitrary manner. Often, there is a lack of understanding of such parameters, such as a reproduction number representing a disease’s ability to spread [30], which can lead to ill-informed decisions and public health interventions by different stakeholders. Determining the effectiveness of interventions, by combining reporting and cross-checking with multiple indicators and data sources, is essential. In particular, there is a need to support surveillance practitioners with ad hoc querying over integrated data. Existing infrastructures are limited to delivering information on a fixed set of defined parameters. Agility is essential and surveillance systems relying on slow to deploy information gathering pipelines lacking interoperability are insufficient, particularly when requirements shift e.g. to understanding demographics of infections, in addition to overall infection rates.

In light of these new requirements for surveillance systems, a new generation of surveillance platforms is emerging that can address the provision of agility and ad hoc querying for non-technical users. Surveillance systems need to be able to discover and select data sets that contribute to a given line of inquiry from a registry of available data services and data transformation services. The essential tasks that need to be supported are (i) the use of precise and formal semantics to describe input and output of data services to ensure they are rapidly discoverable at the time of the query, (ii) the provision of search engines that can understand such semantic descriptions, and (iii) the provision of interoperability between services to ensure complex workflows needed for retrieving and transforming data are uninterrupted and (iv) the provision of intelligible query composition tools that are readily explainable to novice users.

For over a decade, these design requirements have been core to semantic web services frameworks which have been recently deployed in surveillance use cases. In [28, 33, 34] the Semantics, Interoperability, and Evolution for Malaria Analytics (SIEMA) platform was deployed for use in malaria surveillance based on semantic data federation. SIEMA’s objective was to address the interoperability between evolving malaria data sources and provide advanced query options [34] for users with little or no technical skills. The platform leverages Semantic Automated Discovery and Integration (SADI) [31] Semantic Web Services and a semantic query engine HYDRA [32] to implement the target queries typical of malaria programs. The platform uses community-developed Malaria ontologies, to describe data services. It enhances the findability of distributed data resources, and the construction of workflows to fetch data from different Web services.

Al-Manir et al. [28] reported on use cases provided by the Uganda Ministry of Health to illustrate effectiveness in providing seamless access to distributed data and preservation of interoperability between online resources. Specifically, the queries investigate the nature of interventions e.g. Which indoor residual spraying used permethrin as an insecticide?, and more complex queries looking at the impacts of interventions e.g. Which districts of Uganda that used permethrin-based long-lasting insecticide-treated nets in 2015 saw a decrease in *Anopheles gambiae* s.s. population but no decrease in new malaria cases between 2015 and 2016? This latter query is a particularly complex query that involves a combination of multitude of services discovered and orchestrated into a workflow by the HYDRA query engine (See Figure 1 for the list of registered services).

**Figure 1.**
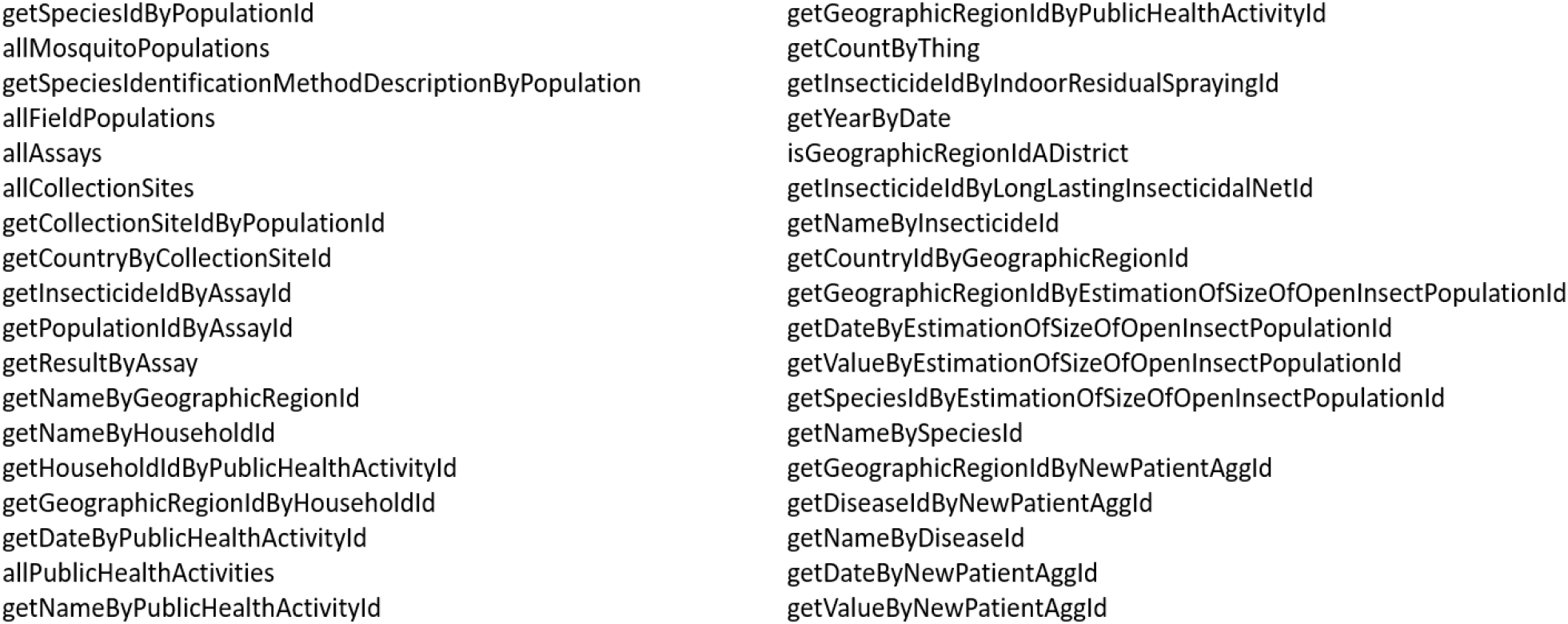
List of registered services.

Figure 2 shows a graphical query presented in the HYDRA GUI and the corresponding SPARQL query. Keyword /graphical inputs presented on a canvas are converted to SPARQL queries and presented to HYDRA for processing. These semantic queries are sharable, editable, and offer a high degree of intelligibility for surveillance experts. There is significant flexibility provided to compose regular and ad hoc queries independent of users needing to understand a data structure or a query syntax. Queries posed to the SIEMA surveillance platform are translated into workflows of services by HYDRA which are composed of one or more SADI services identified in the registry.

**Figure 2.**
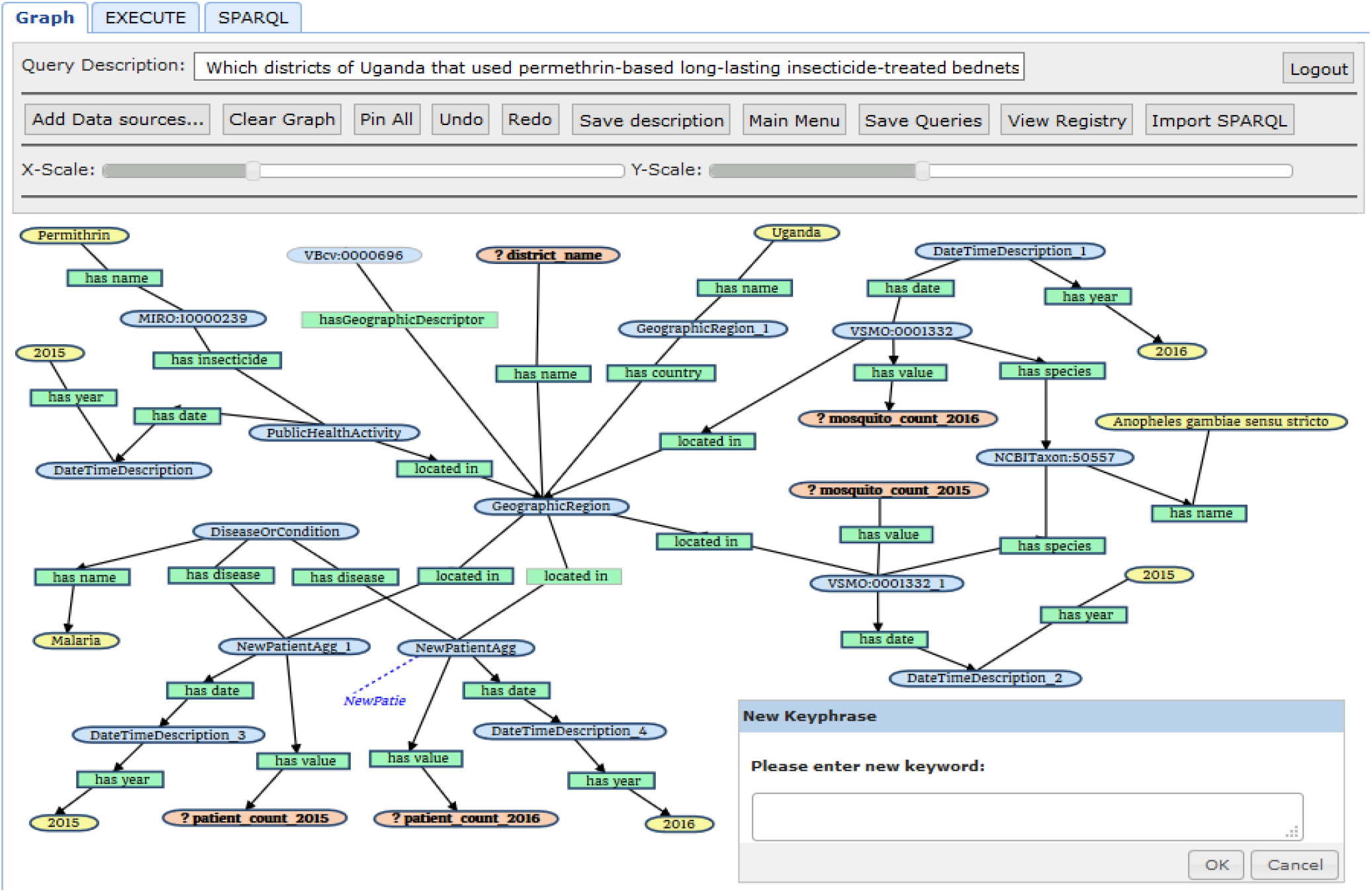
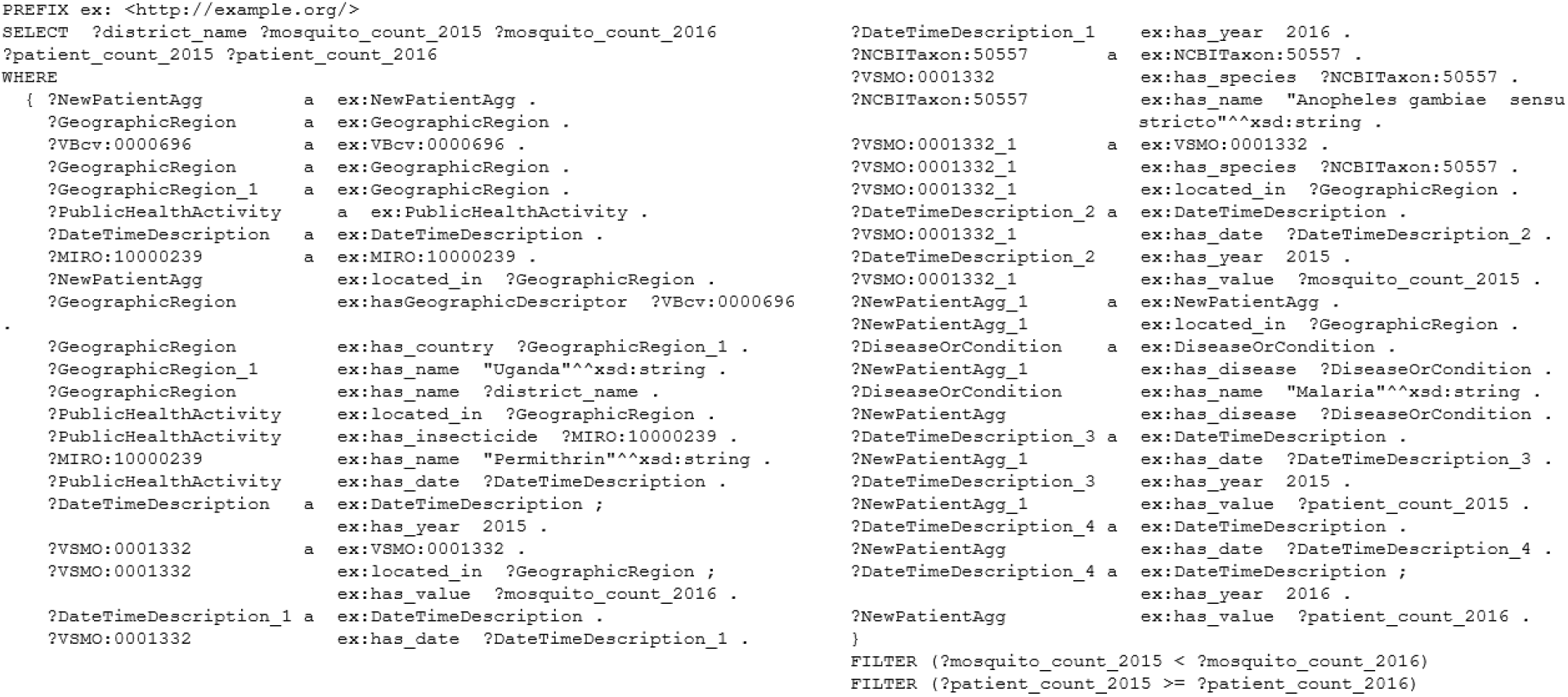
SPARQL query and graphical representation for “Which districts of Uganda that used permethrin-based long-lasting insecticide-treated nets in 2015 saw a decrease in *Anopheles gambiae* s.s. population but no decrease in new malaria cases between 2015 and 2016?”

For explainability, the service interface of each SADI service is based on the myGrid/Moby service Ontology [35] which requires that a service contains information such as its unique name, the URI to locate the service, the URIs where the input and output are defined, and a textual description. Information about the input and output of the service can be explained from the concepts and relations used in their definition. The concepts and relations, which are derived from community adopted standard vocabularies, are all resolvable through their URIs. The services themselves are resolvable from their URIs. Thus, the workflow is explainable as each service and the associated metadata about the input and output of a service is explainable.

The system described in [28, 33] serves as a functioning prototype funded by the Bill and Melinda Gates Foundation Grand Challenges. It demonstrates the state of the art with respect to surveillance. Further deployment of this approach will likely emerge for other surveillance tasks given that the terminology layer and registry used in the framework can be exchanged for other specific terminologies.

## 4 Maintaining Interoperability

Another challenge that is symptomatic of existing surveillance systems is that they can be brittle, in the sense that even minor updates to core terminologies by domain experts, which occur regularly and incrementally, can render them inactive. The challenge is to preserve the integrity and consistency of an integrated system (interoperability). Here the primary activities during this change management activity are detection, representation, validation, traceability, and rollback, as well as the reproduction of the changes [36]. Mitigating this challenge is also a feature of the SIEMA platform [33] which incorporates a custom algorithm to detect changes to community-developed terminologies, data sources, and services and reports these details to a dashboard displaying real-time service availability (uptime/downtime). Based on the type of change detected and its impact on the status of a service, the dashboard is updated. This level of reporting makes it possible for surveillance practitioners to invoke Valet SADI [37] to automatically rebuild services as needed, mitigating service downtime to ensure reliability.

## 5 Conclusion and Outlook

It is clear that pandemic preparedness is an essential theme for the future. Smart surveillance centers and observatories will be required for each municipality and regional governments to archive a range of key datasets. Global Health Observatories [38] have already recorded more than key 1000 indicators for 194 WHO’s member states. Smart cities will need to incorporate provisions for effective interventions, that will need to target both commercial and residential sectors, such as social distancing measures required during pandemics. Primary data, including sensor datasets, need to be readily available for secondary uses in unanticipated ways to improve decision making by governments and civic leaders while ensuring privacy protection and adhering to ethical standards and guidelines

The integration of such datasets will be of paramount importance and modeling of these data for reporting purposes must be prepared in advance. Semantics and ontologies will play an essential role in ensuring interoperability and rapid reuse of data sets for reporting. Preparedness as an activity must include dynamic data sources and all aspects of digital infrastructures that support decision making. In the context of the 2020 Covid-19 pandemic, the Ontology of Coronavirus Infectious Disease (CIDO) [40] was developed to provide standardized human- and computer-interpretable annotation and representation of various coronavirus infectious diseases, including their etiology, transmission, pathogenesis, diagnosis, prevention, and treatment. More specifically the COVID-19 Surveillance Ontology [41] is an application ontology used to support COVID-19 surveillance in primary care. The ontology facilitates monitoring of COVID-19 cases and related respiratory conditions using data from multiple brands of computerized medical record systems. It is anticipated that these ontologies will be expanded by integrating further knowledge from relevant domains to provide a comprehensive semantic backbone necessary for intelligent global pandemic surveillance and policymaking that is essential today.

## Data Availability

All data sources are publicly available.

